# Combined oropharyngeal/nasal swab is equivalent to nasopharyngeal sampling for SARS-CoV-2 diagnostic PCR

**DOI:** 10.1101/2020.06.05.20123745

**Authors:** Tania Desmet, Peter De Paepe, Jerina Boelens, Liselotte Coorevits, Elizaveta Padalko, Stien Vandendriessche, Isabel Leroux-Roels, Annelies Aerssens, Steven Callens, Eva Van Braeckel, Thomas Malfait, Frank Vermassen, Bruno Verhasselt

## Abstract

**Background:** Early 2020, a COVID-19 epidemic became a public health emergency of international concern. To address this pandemic broad testing with an easy, comfortable and reliable testing method is of utmost concern. The nasopharyngeal (NP) swab sampling is the reference method though hampered by international supply shortages. A new oropharyngeal/nasal (OP/N) sampling method was investigated using the more readily available throat swab.

**Methods:** In this prospective observational study 36 COVID-19 patients were tested with both a NP and combined OP/N swab for SARS-CoV-2 RNA by PCR. In hospitalized suspect patients, who tested negative on both swabs, extensive retesting was performed. The sensitivity of NP versus combined OP/N swab sampling on admission and the correlation between viral RNA loads recovered was investigated.

**Results:** 35 patients were diagnosed with SARS-CoV-2 by means of either NP or OP/N sampling. The paired swabs were both positive in 31 patients. The one patient who tested negative on both NP and OP/N swab on admission, was ultimately diagnosed on bronchoalveolar lavage fluid. A strong correlation was found between the viral RNA loads of the paired swabs (r = 0.76; P < 0.05). The sensitivity of NP and OP/N analysis in hospitalized patients (n = 28) was 89.3% and 92.7% respectively.

**Conclusions:** This study demonstrates equivalence of NP and OP/N sampling for detection of SARS-CoV-2 by means of rRT-PCR. Sensitivity of both NP and OP/N sampling is very high in hospitalized patients.

## INTRODUCTION

A pandemic of respiratory disease caused by SARS-CoV-2 began in Wuhan, China in December 2019 and quickly spread to every continent. On February 1^st^, 2020, the disease was declared a Public Health Emergency of International Concern (PHEIC) by the World Health Organization (WHO). (1) By mid-May 2020, over 4 million people were infected and almost 300.000 died.

Real-time reverse transcriptase-polymerase chain reaction (rRT-PCR) on nasopharyngeal swab material is typically used to confirm the clinical diagnosis. (1,2) Nasopharyngeal (NP) swab remains the reference sampling method, while recent studies suggest that nasal swab and saliva sampling may be nearly equivalent. (https://www.who.int/publications-detail/laboratory-testing-for-2019-novel-coronavirus-in-suspected-human-cases-20200117; https://www.cdc.gov/coronavirus/2019-nCoV/lab/guidelines-clinical-specimens.html; 5–6) The detection of SARS-CoV-2 in oropharyngeal swabs seems less sensitive. (7–12)

In order to address a pandemic, extensive mapping and therefore broad testing with an easy and comfortable sampling method is of utmost importance. In the context of an international shortage in nasopharyngeal swabs, the search for a suitable alternative is a global health priority. This study evaluates a new combined oropharyngeal/nasal (OP/N) sampling method using the more readily available throat swab.

## METHODOLOGY

This prospective observational study was conducted at Ghent University Hospital, Belgium from April 7th to May 1st, 2020. Upon presentation at the emergency department, all patients with suspected COVID-19 (based on respiratory and inflammatory symptoms) in whom a NP test was indicated for diagnosis, were candidates for a combined OP/N swab, sampled according to a specific protocol, developed in Ghent University Hospital. At inclusion, no distinction was made based upon disease severity or the need for hospitalization. Hospitalised patients with highly suspect clinical and radiological features of COVID-19 and negative PCR-sample on admission were retested with an additional NP and/or anal swab and finally, if still tested negative, on bronchoalveolar lavage (BAL) fluid for definite PCR-diagnosis. In ambulatory patients no retesting was performed.

Specimens from OP/N were obtained by rubbing the oropharyngeal space twice at both sides of the uvula and placing the same swab into both nasal cavities until a slight resistance was felt (supposed midturbinate). NP sampling was performed after OP/N sampling in order not to ‘contaminate’ the midturbinate part of the nose by viral material from the nasopharynx as may be expected when performed in reversed order. Each swab was rotated three complete turns for optimal mucosal contact. For NP sampling a flexible mini tip flocked swab (in Amies transport medium or in universal transport medium (UTM)) was used (Cat. numbers 481CE and 305C respectively, Copan^®^, Italy). OP/N sampling was performed using a non-flexible flocked swab with normal tip (in Amies transport medium, Cat. number 480CE, Copan^®^). rRTPCR for SARS-CoV-2 RNA was performed on both the NP swab and the OP/N swab according to Corman et al. (13) using NucliSens easyMag™ RNA extraction (bioMérieux, Marcy-l’Étoile, France) and one-step rRT-PCR (Qiagen One Step RT-PCR Kit, Cat. number 210212, Qiagen, Hilden, Germany) on CFX96 cyclers (Bio-Rad, Hercules, USA). All tests were performed in-hospital on a daily basis. A cycle threshold (Ct) value below 50 was interpreted as positive for SARS-CoV-2 RNA.

All clinical samples were tested for inhibition by adding 7.5 µl of Diagenode RNA extraction and inhibition Real-Time PCR control (Cat. Number DECR-CY-L100, Diagenode SA^®^, Belgium).

Since retesting was not performed in the ambulant patient group, sensitivity of OP/N and NP sampling was considered in hospitalized patients only.

Descriptive and relative frequencies were used to describe the distribution of cases. Since the data were paired, the Wilcoxon signed-rank test was used to assess differences between cycle threshold (Ct, considered as log2 transformed data of quantity viral RNA load) in NP and OP/N specimens. The correlation between NP PCR Ct and OP/N PCR Ct was assessed using Spearman’s rank correlation. Sensitivity was calculated by means of cross tabs. Specificity was found not to be relevant since not one false positive SARS-CoV-2 PCR-result was found in this trial period, in line with literature. (5) Due to the nature of this trial, assessing sensitivity without a negative control group, a calculation of the weighted kappa coefficient was not relevant. All analyses were performed using IMB^®^ SPSS^®^ Statistics version 26.

The study was approved by the ethical review board Ghent University Hospital (BC-07662).

## RESULTS

During the study period, 41 SARS-CoV-2 PCR-confirmed patients were diagnosed at Ghent University Hospital. Five patients were excluded because no OP/N swab was sampled upon admission. Seventy-five samples from the 36 remaining patients were analysed. The median age was 61 years (range, 22-90), 21 out of 36 patients (58%) were male and 28 (78%) patients were hospitalized. Overall, 35/36 patients were diagnosed with SARS-CoV-2 by means of either NP or OP/N sampling on admission. The paired swabs were both positive in 31 patients. In two patients, NP swabs were positive (Ct 36.47 and 38.38) with negative OP/N swabs. In two other patients, OP/N swabs were positive (Ct 34.14 and 41.39) with negative NP swabs. The one patient who tested negative on both NP and OP/N swab at admission, was ultimately diagnosed on BAL fluid (Ct 35.80).

When the paired OP/N and NP samples were compared in the 31 patients who tested positive in both, the median Ct for SARS-CoV-2 PCR was not significantly different: 27.5 cycles (range, 15.7–40.4) in NP samples vs. 27.2 cycles (range, 14.1–40.2) (P = 0.576) in OP/N samples.

Besides, a strong correlation was found between the Ct values of the paired swabs (r = 0.76; P < 0.05) (Figure 1). From the 28 hospitalized patients, 27 were diagnosed by means of either NP (25) or OP/N (26) sampling. Thus, when considering a PCR positive result on BAL fluid as the ultimate confirmation of COVID-19 diagnosis, sensitivity of NP and OP/N analysis were 89.3% and 92.7% respectively in hospitalized patients (n = 28).

**Figure 1.**
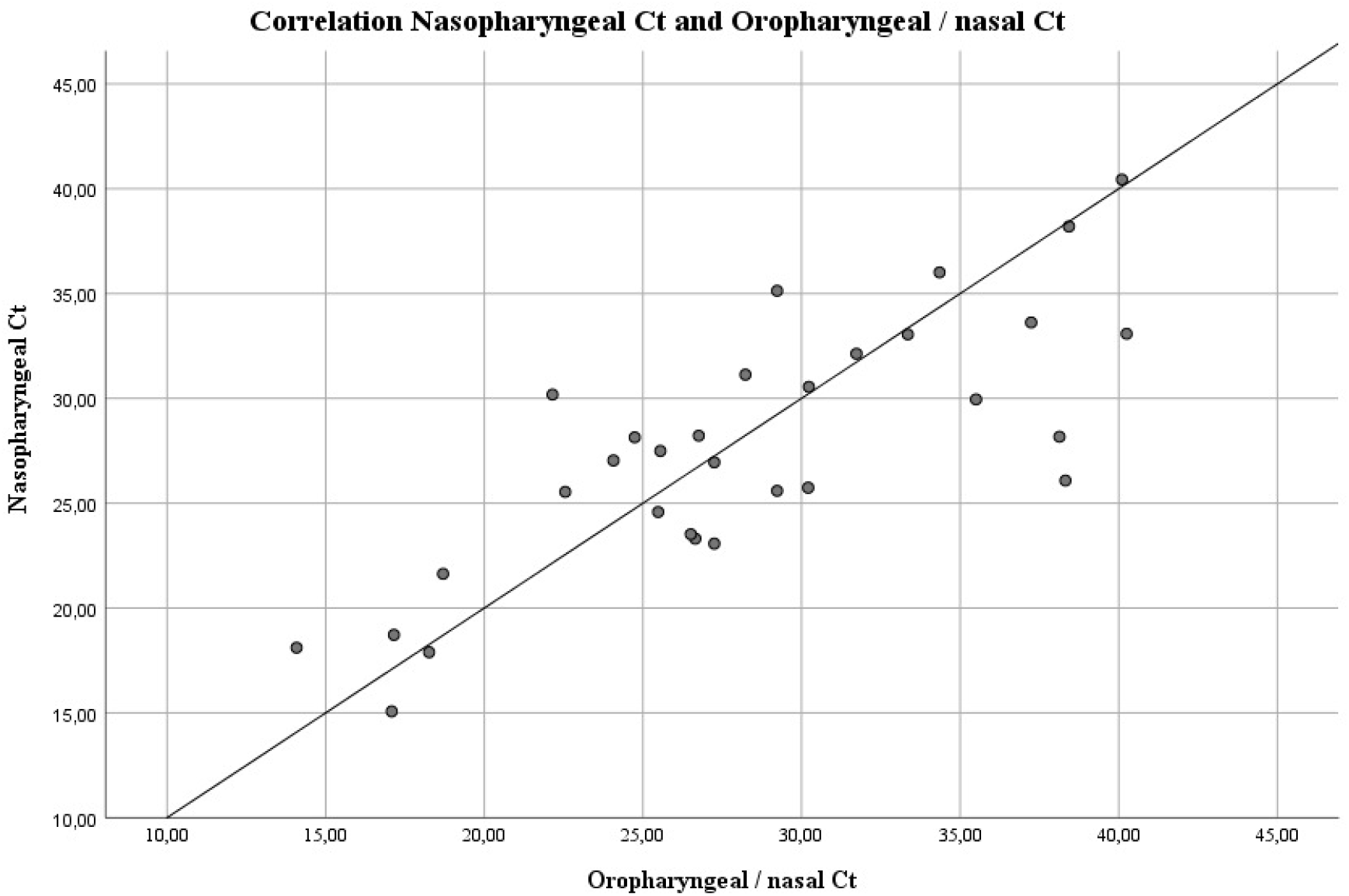
Correlation of viral RNA load (log2 transformed as threshold cycles) in nasopharyngeal sampling and oropharyngeal/nasal sampling. R = 0.76.

## DISCUSSION

Nasopharyngeal swab is considered to be the reference sampling method in suspected COVID-19 patients. (https://www.who.int/publications-detail/laboratory-testing-for-2019-novel-coronavirus-in-suspected-human-cases-20200117, https://www.cdc.gov/coronavirus/2019-nCoV/lab/guidelines-clinical-specimens.html) In the context of an international shortage in nasopharyngeal swabs, the search for a suitable alternative is a burning issue. Moreover, given the need of extensive population testing, an easy and patient-friendly sampling method is preferred. Salivary sampling has been reported as a reliable tool to detect SARS-CoV-2 in COVID-19 patients. This study, however, did only include patients with severe to very severe disease and did not provide information on mildly or moderately ill patients. (6) Nasal sampling was proposed as an alternative to nasopharyngeal sampling, though appears to be inferior to nasopharyngeal sampling (9.1% false negatives). (5) Oropharyngeal (OP) sampling is less sensitive (compared both to nasopharyngeal and nasal sampling) according to several studies. Wang X. et al. (2020) and Wang W. et al. (2020) reported a positivity rate of OP sampling of 27% and 32% respectively in COVID-19 patients. Sensitivity results were not reported.(8,11)

To our knowledge, our study is the first to show that the molecular detection of SARS-CoV-2 on combined OP/N swabs was non-inferior to nasopharyngeal sampling. (https://www.who.int/publications-detail/laboratory-testing-for-2019-novel-coronavirus-in-suspected-human-cases-20200117, https://www.cdc.gov/coronavirus/2019-nCoV/lab/guidelines-clinical-specimens.html) We found a high correlation between the viral RNA loads of the paired OP/N and NP samples. Besides, the sensitivities of both NP and OP/N sampling in hospitalized patients were 89.3% and 92.7% respectively. Large differences in sensitivity of NP sampling were published ranging from 63% to 73.3%. (Y. Yang-unpublished data, 11,14) It should be mentioned however that, to our knowledge, no peer-reviewed studies or large trials regarding this subject are available. The gold standard to which tests under investigation are to be compared, is not clear and differs among various publications, although consensus seems to exist about the positioning of BAL fluid as the most reliable testing method. (Y. Yang, unpublished data)

This study, performed in symptomatic patients with a broad age range and various degrees of disease severity, demonstrates equivalence of NP and OP/N sampling for detection of SARS-CoV-2 by means of rRT-PCR. Sensitivity of both NP and OP/N sampling is very high in hospitalized patients. These results warrant further evaluation of the combined oropharyngeal/nasal testing strategy in pauci- or asymptomatic individuals in order to tackle the global shortage in NP swabs. In this context, the new sampling strategy described here was used by the Belgian Government to guide a national screening campaign in residential care centres.

## Data Availability

Raw data are available upon request

## ACKNOWLEDGEMENTS

The authors do not have an association that might pose a conflict of interest.

Desmet Tania: No conflict.

De Paepe Peter: No conflict.

Boelens Jerina: No conflict.

Coorevits Liselotte: No conflict.

Padalko Elizaveta: No conflict.

Vandendriessche Stien: No conflict.

Leroux-Roels Isabel: No conflict.

Aerssens Annelies: No conflict.

Van Braeckel Eva: No conflict.

Malfait Thomas: No conflict.

Callens Steven: No conflict.

Vermassen Frank: No conflict.

Verhasselt Bruno: No conflict.

